# History of coronary heart disease increases the mortality rate of COVID-19 patients: a nested case-control study

**DOI:** 10.1101/2020.03.23.20041848

**Authors:** Tian Gu, Qiao Chu, Zhangsheng Yu, Botao Fa, Anqi Li, Lei Xu, Yaping He, Ruijun Wu

## Abstract

**Background:** China has experienced an outbreak of a novel human coronavirus (SARS-CoV-2) since December 2019, which quickly became a worldwide pandemic in early 2020. There is limited evidence on the mortality risk effect of pre-existing comorbidities for coronavirus disease 2019 (COVID-19), which has important implications for early treatment.

**Objective:** Evaluate the risk of pre-existing comorbidities on COVID-19 mortality, and provide clinical suggestions accordingly.

**Method:** This study used a nested case-control design. A total of 94 publicly reported deaths in locations outside of Hubei Province, China, between December 18^th^, 2019 and March 8^th^, 2020 were included as cases. Each case was matched with up to three controls, based on gender and age ± 1 year old (94 cases and 181 controls). The inverse probability weighted Cox proportional hazard model was performed.

**Results:** History of comorbidities significantly increased the death risk of COVID-19: one additional pre-existing comorbidity led to an estimated 40% higher risk of death (p<0.001). The estimated mortality risk in patients with CHD was three times of those without CHD (p<0.001). The estimated 30-day survival probability for a profile patient with pre-existing CHD (65-year-old female with no other comorbidities) was 0.53 (95% CI [0.34-0.82]), while it was 0.85 (95% CI [0.79-0.91]) for those without CHD. Older age was also associated with increased death risk: every 5-year increase in age was associated with a 20% increased risk of mortality (p<0.001).

**Conclusion:** Extra care and early medical intervention are needed for patients with pre-existing comorbidities, especially CHD.

## Introduction

Since the first report of Coronavirus Disease 2019 (COVID-19) in December 2019 in Wuhan, Hubei Province, China, the novel virus infection has rapidly spread to other cities in China, and has now been detected in 186 countries and locations internationally [1]. On March 11^th^, 2020, the World Health Organization declared COVID-19 a pandemic, and has called for aggressive actions from all countries to fight the disease. Current research has indicated that COVID-19 is caused by severe acute respiratory syndrome coronavirus 2 (SARS-CoV-2), a beta-coronavirus similar to the severe acute respiratory syndrome-coronavirus (SARS-CoV) in the genetic sequence [2]. Epidemiological evidence suggests that initially reported cases in China had a history of exposure to the Huanan seafood market [3-4]. With the escalated spread of the infection, there has been clear evidence of human to human transmission [5-6]. The most common symptoms include fever, dry cough and fatigue [5-8], with presence of asymptomatic, yet contagious cases [9].

According to the COVID-19 situation reports of WHO, as of March 29^th^, 2020, the infection has caused 81,445 confirmed cases in mainland China, including 3,300 deaths. Internationally, a total of 601,020 confirmed cases have been reported from 186 countries outside of China, including 28,747 deaths. Considering the global public health threat posed by COVID-19, unraveling the prognostic factors for patients, especially the risk factors of mortality associated with COVID-19, has important implications for clinical practice and is urgently warranted.

Studies have indicated that severe cases tend to be older in age [6, 8] and are more likely to have had pre-existing medical conditions, including but not limited to hypertension [3, 6, 8], diabetes [6, 8], cardiovascular diseases [3, 6, 8, 10-12], cerebrovascular diseases [6], chronic obstructive pulmonary disease (COPD) [3, 8], cancer [13], and digestive diseases [14], in comparison to non-severe cases [3, 6, 8-15]

Recently, scholarly attention focuses on identifying the risk factors for death from COVID-19. Some evidence suggests that pre-existing medical conditions are likely death risk factors for COVID-19. For example, a study based on 72,314 cases in China indicated that the case-fatality rate (CFR) tends to be higher among those with older age, and having pre-existing cardiovascular disease, diabetes, and hypertension compared to all the patients [9]. Similarly, by conducting logistic regression on odds of in-hospital death among 54 diseased patients and 137 recovered patients in Wuhan City, Hubei Province, Zhou et al. [16] found that older age, higher Sequential Organ Failure Assessment (SOFA) score and d-dimer greater than 1ug/ml at hospital admission were associated with increased odds of in-hospital death. Chen et al. (2020) found that pre-existing hypertension and other cardiovascular complications were more common among diseased patients than recovered patients (unpublished data).

However, several gaps remain in the understanding of risk factors for mortality of COVID-19. First, most current research on pre-existing comorbidities of COVID-19 was based on univariate comparison, which did not account for important confounders such as age and gender [17-20]. Second, no studies have investigated the hazard of the identified risk factors over time, or the probability of survival at a given time. Under the rapidly changing pandemic situation, it is crucial to provide timely survival-time guidance for implementing the targeted treatment to the high-risk patients in clinical practice. Third, most existing studies on mortality risk factors were focused on patients diagnosed in Wuhan, Hubei Province, with little understanding about the mortality risk factors outside of Hubei Province. The risk factors are likely different inside and outside of Hubei Province, since current research has found that the clinical symptom severity [5] and the fatality-case rate [9, 21] to be higher in Hubei Province (the center of outbreak) than cities outside of Hubei Province in China. Fourth, no studies thus far have taken into account the pandemic stage when evaluating mortality risk factors. It has been found that average daily attack rate in China was different before and after January 11^th^ 2020, since non-pharmaceutical interventions were taken by the government before this date [22]. The change of pandemic stage may also influence the risk factors for fatality associated with COVID-19.

To fill the above research gaps in the existing literature about the morality risk factors for COVID-19, the present study conducted a nested case-control (NCC) study, aiming to evaluate the risk of the common pre-existing comorbidities (hypertension, coronary heart disease, diabetes and etc.) for mortality associated with COVID-19 in mainland China outside of Hubei Province. NCC, also called risk set sampling, has been widely used in studying the fatal disease risk effect in large pharmacoepidemiologic studies [23-28] and risk prediction in pandemic influenza A (H1N1) 2009 (pH1N1) [29]. NCC is cost-effective in data collection, and is especially suitable for research on the death risk of diseases such as COVID-19, where the number of event-free people largely exceeds those who experienced events [30]. To attain this goal, we employed survival analysis on 275 publicly reported confirmed cases, adjusting for age, gender and the change in the pandemic stage in China (i.e., before and after January 11^th^, 2020).

## Method

### Study Design and Rationale

This study performed survival analysis under a nested case-control (NCC) design to assess the roles of common comorbidities (cardiocerebrovascular, endocrine and respiratory disease, etc.) in predicting mortality for COVID-19, among patients in mainland China outside of Hubei Province. The study period was from December 18^th^, 2019, when the first laboratory-confirmed case was announced in China, till March 8^th^, 2020.

The study cohort was defined as all the publicly reported confirmed COVID-19 patients outside of Hubei Province in mainland China between the study period. During this period, 112 deaths outside of Hubei Province were reported by the National Health Committee of China, and 18 were excluded from the present study due to missingness of important clinical information. A total of 448 publicly reported laboratory-confirmed COVID-19 cases (94 deaths and 354 survivors) were initially collected. The data collection procedure was blinded to patient comorbidity information. All deaths were included as cases, and each case was matched with up to three controls on gender and age ±1 year old (94 cases and 181 controls). The sample distribution across all 32 province-level regions in mainland China is presented in Appendix Table A1.

### Data Collection Procedure

We routinely searched for daily news and public health reports on confirmed COVID-19 cases in all areas in mainland China outside of Hubei Province. Patients’ clinical and comorbidity characteristics were recorded and doubly confirmed by national/provincial/municipal health commission websites, the official COVID-19 data reporting websites in China. Follow-up time was defined as the duration from the date of disease onset till the end of observation on March 8^th^ or when the participant died, whichever came first. For each eligible patient, we followed local reports to update their survival status until the end of follow-up time.

As illustrated in Figure 1, the inclusion criterion was publicly reported COVID-19 patients who had complete information on basic demographics (age, gender and region), disease onset date--the first time a patient became symptomatic, and history of comorbidities (include but not limited to hypertension, cardiovascular disease, diabetes and respiratory diseases) were included in the analysis. Asymptomatic patients were not included in this study. In addition, we defined “comorbidity-free patients” as those who were specifically described as “no pre-existing medical condition/comorbidity” on the national/provincial/municipal health commission websites.

**Figure 1:**
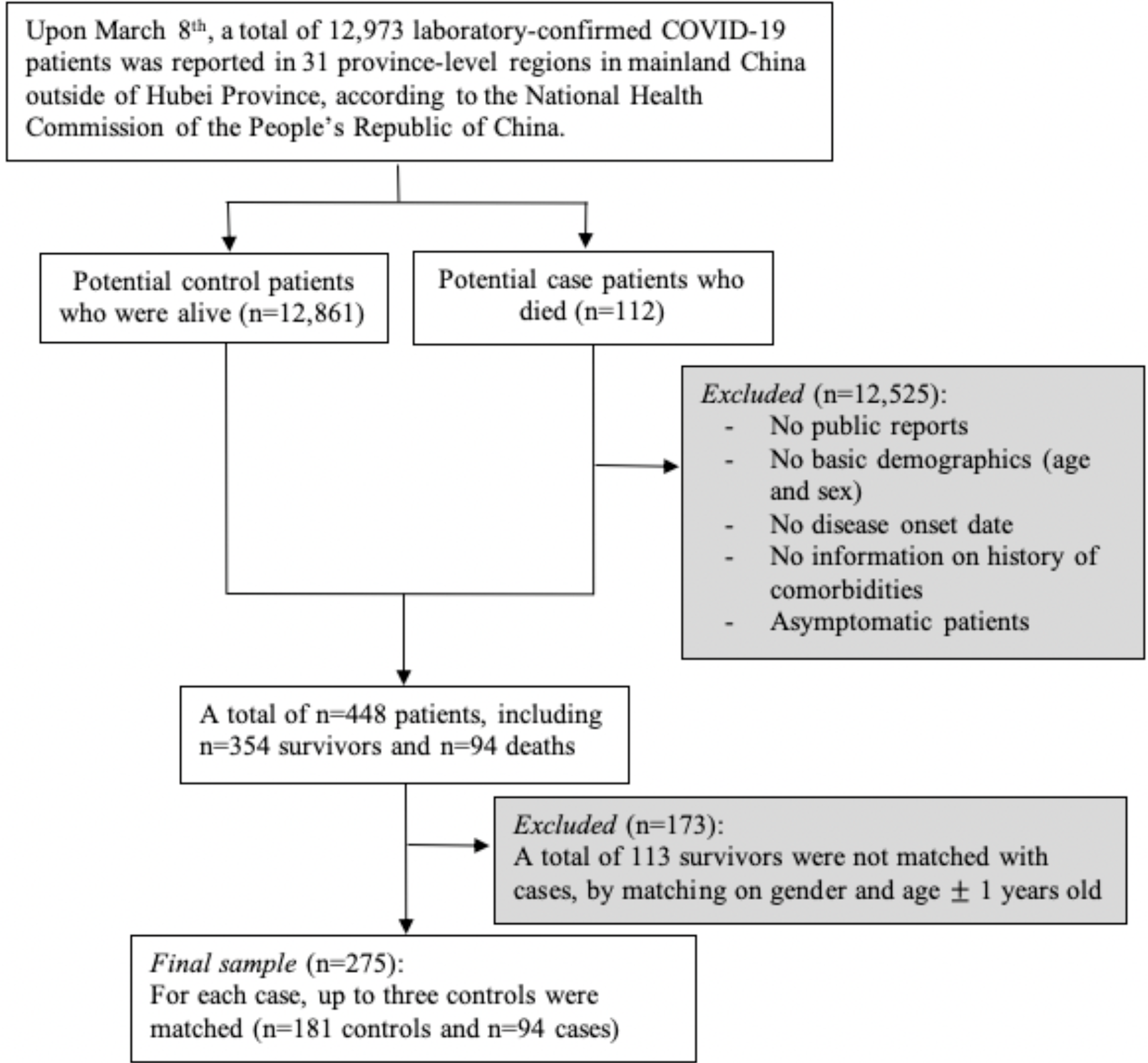
Patient flow diagram detailing included subjects and exclusion criteria

In the following three steps, we used the No. 261 patient in the sample as an example to introduce the dynamic tracking method we used to identify any missing dates:

#### Step 1

Conducting an internet search on confirmed cases on baidu.com, the largest search engine in China, using keywords “confirmed COVID-19 cases report” and “pre-existing comorbidities.” A search result pertained to one confirmed case reported on the website of Municipal Health Commission of Binzhou (Shandong Province) on February 17^th^, described as “*the 15*^*th*^ *confirmed case: 30-year-old male without pre-existing morbidities, who lives in the neighborhood of Xincun Village. This patient was diagnosed positive on February 16*^*th*^ *and is being treated with precaution in Bincheng hospital*.*”* We recorded age, gender, region and comorbidity-free for this patient.

#### Step 2

We then determined the onset date of this patient based on another announcement on the same website. In this announcement titled “*Possible exposure locations and times of the 15*^*th*^ *confirmed case*,” it says, “*the patient was symptomatic on February 14*^*th*^.*”*

#### Step 3

Finally, we confirmed the event status of this patient as discharged on March 3^rd^, by following the updates on this website.

### Statistical Analysis

Analyses were performed in R 3.6.2 (R Foundation for Statistical Computing). Baseline clinical characteristics were shown as mean (SD), median (range), or number (%), with a comparison of characteristics in subjects stratified by case and control via t-test for continuous variables and chi-squared or Fisher’s exact test for binary variables.

In order to utilize the time-to-event information under the NCC design, the inverse probability weighting Cox proportional hazard regression model was employed [32]. The matching between cases and controls, and relative weights were simultaneously obtained via KMprob function in multipleNCC R package [31], by specifying the Kaplan-Meier type weights with additional matching on gender and age ±1 year old. Only survivors were assigned weights, since all cases (deaths) were included as designed with a weight of one. Those survivors with sampling probabilities of zero were considered as “fail to match” and excluded from the study.

The total number of comorbidities was defined as the summation of comorbidities, ranging from zero to four or above. Kaplan-Meier curve was plotted to check the proportional hazard assumption and the Pearson correlation test was used to rule out the multicollinearity concern before fitting any model. Univariate weighted Cox models were performed for each comorbidity. The multivariate weighted Cox model was used to determine if pre-existing comorbidity yielded prognostic hazard information. We included those comorbidities that were marginally significant (p<0.1) in the univariate analysis to the multivariate model. Other than the common risk factors (age and gender), the multivariate model also adjusted for early period of pandemic (after vs. before January 11^th^, 2020 when no-intervention was taken by the government) [22]. Although matching was based on age and gender, we adjusted for the matching covariates, since the matching was broken with inverse probability weighting [32]. A separate multivariate model was built by using the total number of pre-existing comorbidities as an ordinal predictor, adjusting for the same covariates. Hazard ratios (HRs) from the weighted Cox model were reported along with 95% confidence intervals (CIs) and p-values. Sensitivity analysis was performed using multivariate logistic regression to provide estimated odds ratio (ORs), which includes the same covariates as the multivariate weighted Cox model.

Weighted Cox model-based survival estimates were plotted for an example patient profile (65-year-old female with no other comorbidities) to compare the survival probability over time with and without CHD. The log-rank test was used to compare the median survival difference.

### Ethics Approval

The study was approved by Shanghai Jiao Tong University Public Health and Nursing Medical Research Ethics Committee (SJUPN-202001).

## Results

### Sample Description

Table 1 summarizes patient demographics and pre-existing medical conditions. Results are presented for all patients in the study (n=275), as well as for cases (n=94) and controls (n=181), respectively. Patients were 24-94 years old (Mean_age_ = 66.4, SD_age_ =14.5). The average age tended to be older in the case group (70.7 years old) than in the control group (64.2 years old). Median ages were similar to mean ages in both groups. A majority (62.9%) of the patients were male. Overall, 25.5% of the total patients had clinical symptoms associated with COVID-19 before January 11^th^, 2020. A relatively small proportion of the total sample had COPD, renal failure, history of surgery and hepatic failure (4.4%, 4.4%, 3.6% or 1.1%). Among all pre-existing comorbidities with over 5% of the total sample, hypertension was the most common (39.6%), followed by diabetes (26.2%), CHD (14.5%), cardiac failure (8%), cerebral infarction (6.9%) and chronic bronchitis (6.9%). Patients in the case group had more CHD (p<0.001) and more cerebral infarction (p=0.05), compared to those in the control group. A majority of the total sample had at least one pre-existing comorbidity (67.6%), specifically around 25% had one, 22% had two, 11% had three, and 10% had four or more. Compared to the control group, more patients in the case group had pre-existing comorbidities (p=0.02), especially those who had four or more comorbidities (p<0.001).

**Table 1.** Patient Characteristics, stratified by survival status*

|  | Overall<br>(N=275), n (%) | Case (deaths)<br>(N=94), n (%) | Control (survivors)<br>(N=181), n (%) | P Value† |
| --- | --- | --- | --- | --- |
| **Matching variables** |  |  |  |  |
| Age |  |  |  |  |
| Mean (SD) | 66.4 (14.5) | 70.7 (13.3) | 64.2 (14.7) | **<0.001** |
| Median (Min, Max) | 68.0 [24.0, 94.0] | 72.5 [25.0, 94.0] | 67.0 [24.0, 90.0] | - |
| Male | 173 (62.9) | 56 (59.6) | 117 (64.6) | 0.49 |
| **Other covariates** |  |  |  |  |
| Before 01/11/2020 | 70 (25.5) | 27 (28.7%) | 43 (23.8) | 0.52 |
| History of surgery | 10 (3.6) | 4 (4.3) | 6 (3.3) | 0.74 |
| **Cardiocerebrovascular diseases** |  |  |  |  |
| Hypertension | 109 (39.6) | 42 (44.7) | 67 (37.0) | 0.27 |
| CHD | 40 (14.5) | 25 (26.6) | 15 (8.3) | **<0.001** |
| Cardiac failure | 22 (8.0) | 10 (10.6) | 12 (6.6) | 0.35 |
| Cerebral infarction | 19 (6.9) | 11 (11.7) | 8 (4.4) | **0.05** |
| **Endocrine diseases** |  |  |  |  |
| Diabetes | 72 (26.2) | 26 (25.4) | 46 (27.7) | 0.80 |
| **Respiratory diseases** |  |  |  |  |
| Chronic bronchitis | 19 (6.9) | 7 (7.4) | 12 (6.6) | 1.00 |
| COPD | 12 (4.4) | 7 (7.4) | 5 (2.8) | 0.14 |
| **Other diseases** |  |  |  |  |
| Renal failure | 12 (4.4) | 6 (6.4) | 6 (3.3) | 0.38 |
| Hepatic failure | 3 (1.1) | 3 (3.2) | 0 (0) | 0.07 |
| **Total number of comorbidities** |  |  |  |  |
| 0 | 89 (32.4) | 21 (22.3) | 68 (37.6) | **0.02** |
| 1 | 68 (24.7) | 18 (19.1) | 50 (27.6) | 0.16 |
| 2 | 61 (22.2) | 22 (23.4) | 39 (21.5) | 0.84 |
| 3 | 30 (10.9) | 14 (14.9) | 16 (8.8) | 0.19 |
| 4 and above | 27 (9.8) | 19 (20.2) | 8 (4.4) | **<0.001** |

CHD: coronary heart disease;
COPD: chronic obstructive pulmonary disease.
\*Mean (standard deviation) is reported for the continuous variables and the counts (%) for categorical variables. p values were calculated by t test, $\chi^2$ test, or Fisher's exact test, as appropriate.
Bold: statistically significant using threshold $p < 0.05$ .

### Model Results

Results of the Pearson correlation test showed no significant correlations among the presence of comorbidities of interest, and the assumption of the proportional hazard was not violated.

Table 2 presents the results of univariate and multivariate weighted Cox models. Older age was associated with significantly higher death risk with similar magnitude in univariate and multivariate models. In the adjusted model, every 5-year increase in age was associated with an estimated 20% higher risk of death (p<0.001). No significant hazard difference was found between male and female patients. Disease infection during the early no-intervention period was associated with a higher risk of death, although not statistically significant.

**Table 2.**
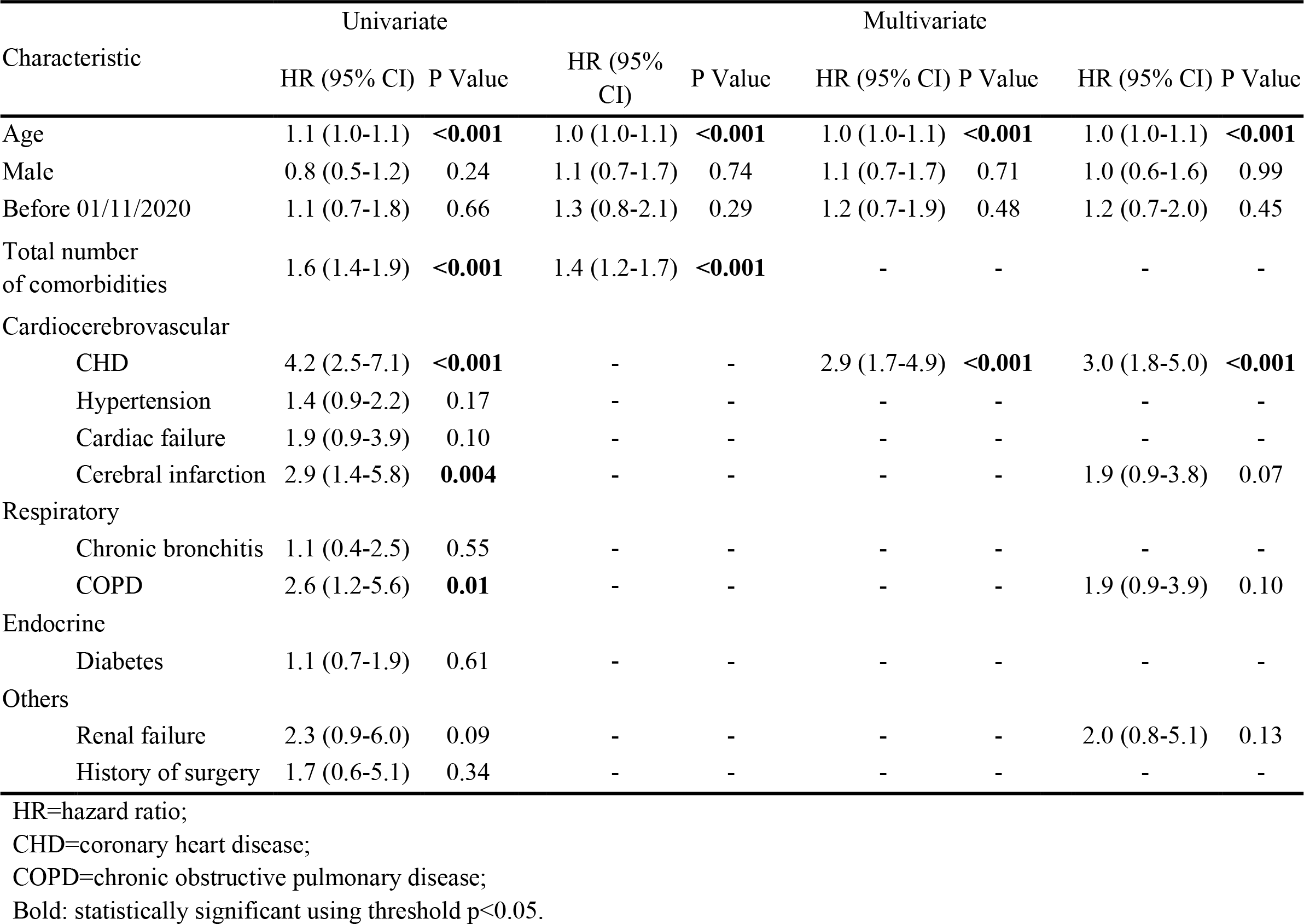
Univariate and multivariate model result from weighted Cox proportional hazard regression

The increasing number of cumulative comorbidities was associated with higher mortality risk in both unadjusted and adjusted models (p<0.001). Moreover, one additional pre-existing comorbidity was associated with a 40% increased risk in mortality of COVID-19. All pre-existing comorbidities were associated with a higher risk of COVID-19 mortality in the univariate model, of which CHD had the largest hazard ratio (HR) of 4.2 (p<0.001), followed by cerebral infarction (HR=2.9, p=0.004), COPD (HR=2.6, p=0.01), renal failure (HR=2.3, p=0.09), cardiac failure (HR=1.9, p=0.1), history of surgery (HR=1.7, p=0.34), hypertension (HR=1.4, p=0.17), diabetes (HR=1.1, p=0.61) and chronic bronchitis (HR=1.1, p=0.55). After adjusting for age, gender, and early period of pandemic in China, CHD was the only comorbidity that yielded a significant death risk: COVID-19 patients with pre-existing CHD had an estimated 2.9 times death risk of those without CHD. In addition, cerebral infarction, COPD, and renal failure all had an estimated HR of around 2.0, respectively. Similar results were observed by using unweighted logistic regression in a sensitivity analysis (Appendix Table A2).

The overall median follow-up was 40 days, during which 94 deaths were observed. Figure 2 shows the estimated survival probability over 70 days for an example patient profile with and without CHD (65-year-old female with no other comorbidities). For such patient profile, having pre-existing CHD led to a significantly shorter survival probability over time, compared to those without CHD (p<0.001). For those with CHD, the estimated 30-day survival probability was 0.53 (95% CI [0.34-0.82]), with an estimated 34 days’ median survival time. On the other hand, the estimated 30-day probability was 0.85 (95% CI [0.79-0.91]) for those without CHD, with the corresponding median survival time over 70 days.

**Figure 2:**
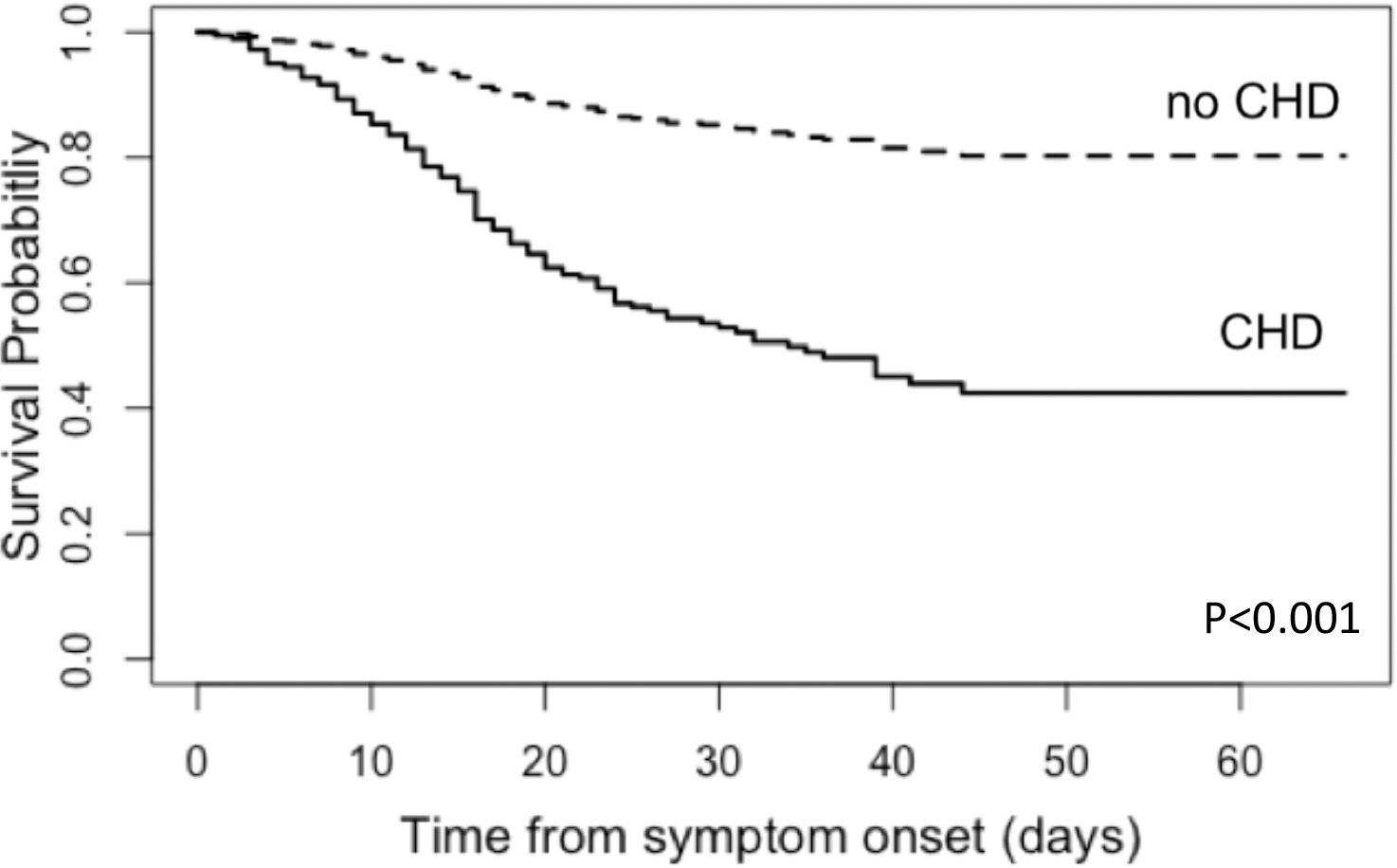
Estimated survival probability over time from the adjusted Cox proportional hazard model for an example patient profile (65-year-old female without CHD [dashed line] or with CHD [solid line], who had no other comorbidities). The estimated 30-day survival probability was 0.53 (95% CI [0.34-0.82]) for patients with pre-existing CHD, while 0.85 (95% CI [0.79-0.91]) for those without (p<0.001).

## Discussion

In this paper, we used survival analysis to estimate the fatal risk of pre-existing comorbidities in COVID-19, based on publicly reported confirmed cases and adjusted for the confounding effect of age, gender, and early period of the pandemic when no-intervention was taken. There were three major findings: First, a history of comorbidities significantly increased the death risk of COVID-19: one additional pre-existing comorbidity led to an estimated 40% increase of death risk (p<0.001). Second, after adjusting for confounders, CHD was the only significant risk factor for COVID-19 mortality. Patients with CHD had a 200% higher risk of mortality in COVID-19, compared to patients without CHD (p<0.001). For a patient profile (65-year-old female without other comorbidities), the estimated median survival times for those with CHD was 34 days, while it was over 70 days for those without CHD (p<0.001). Third, older age was associated with increased death risk. Specifically, every 5-year increase in age was associated with a 20% increased risk of mortality (p<0.001).

To our best knowledge, the present study is the first to provide substantial statistical evidence to show the effect of CHD in predicting mortality for COVID-19. This result is consistent with previous studies that found higher case-fatality rate among patients with cardiovascular disease [9, 15]. It is worth pointing out that the existing studies [9, 16] that investigate prognostic factors for death used chi-square tests or univariate logistic regression that does not control for potential confounders. In contrast, by conducting weighted Cox proportional hazard regression models, our study used time-to-event outcomes, which offers more survival-time-related information to help guide the clinical intervention and more statistical power to detect risk factors [33-34].

Previous studies have indicated that cardiovascular events following pneumonia may increase the risk of mortality [35-41], which explained our findings from the point view of pathophysiological mechanisms. One potential mechanism underlying the association between pneumonia and cardiovascular events is inflammation [38]. Specifically, the inflammatory reaction following pneumonia can result in plague instability and damage in the blood vessels, where evidence of elevated local inflammation in the atherosclerotic coronary arteries following acute systemic infections have been shown in many studies [38, 40]. Thus, infections may result in heightened loading imposed on cardiomyocytes, and lead to sympathetic hyperactivity, ischemia, which may increase the risk of arrhythmia and heart failure in COVID-19 patients with pre-existing CHD [37].

Given the limited understanding of the prognostic factors for COVID-19, more research, potentially prospective studies, are needed to investigate the mechanism by which pre-existing CHD may influence the survival probability among patients with COVID-19. From the clinical point of view, early evaluation of patient medical history is necessary to implement early medical interventions and decrease the mortality risk. We suggest monitoring the dynamic heart rate for patients with pre-existing CHD. For those severe-symptomatic patients who had pre-existing heart ischemia and abnormal heart function, early medical intervention may be needed [41].

Furthermore, our results indicated that the hazard of COVID-19 death was significantly higher in patients at older ages. Adjusting for others, every 5-year increase in age was associated with a 22% increased risk of death, similarly to what was found in previous studies [8, 9, 16]. Alongside the evidence of prognostic risk in CHD, we suggest that extra care is needed for those with CHD, especially for elderly patients.

The design of excluding patients from Hubei Province was based on the concerns of unknown confounders caused by insufficient medical resources in the epicenter. CDC’s report has pointed out that the rapidly increasing number of infections could easily crash the health care system by exceeding its maximum capacity [42]. Therefore, analyzing patient data outside of Hubei Province can avoid the competing death risk caused by insufficient health care resources, and reveal the true underlying impact of pre-existing comorbidities on COVID-19 mortality [44].

One limitation of the present study lies in the nature of publicly reported data. Researchers have pointed out that severe cases may be over-represented in publicly reported data [45]. Nevertheless, we have managed to reduce the potential bias caused by severe case over-representation, through the appropriate matching between cases and controls in NCC design. The auto-matching procedure via statistical program also prevented the possibility of tendentiously selecting survivors with comorbidity-free history during data collection. In addition, NCC design is favored in our situation where the risk factor data and event of interest can be identified opportunistically from publicly reported confirmed cases [30]. Therefore, NCC was the optimal choice, given the restricted availability of public data.

In conclusion, our findings provided preliminary yet strong evidence supporting the association between pre-existing CHD and mortality risk for patients with COVID-19. Based on our findings, close monitoring, extra care, and early medical intervention are needed for patients with pre-existing CHD, to reduce the mortality risk associated with COVID-19.

## Data Availability

Data and code are all available and in preparation.

## Acknowledgement

This project was funded by the National Natural Science Foundation of China (No. 71874111), Shanghai Municipal Health Bureau Foundation (No. 201740116) and Shanghai Jiao Tong University Scientific and Technological Innovation Funds (YG2020YQ01, YG2020YQ06).

---

**Appendix Table A1.** Sample distribution across all 32 province-level regions in mainland China

| Province | Survivors |  | Deaths |  |
| --- | --- | --- | --- | --- |
|  | National total n | Sample n (%) | National total n | Sample n (%) |
| Guangdong | 1343 | 9 (0.67) | 8 | 8 (100.00) |
| Henan | 1265 | 7 (0.55) | 22 | 17 (77.30) |
| Zhejiang | 1205 | 10 (0.83) | 1 | 1 (100.00) |
| Hunan | 1003 | 15 (1.50) | 4 | 2 (50.00) |
| Anhui | 981 | 9 (0.92) | 6 | 6 (100.00) |
| Jiangxi | 932 | 3 (0.32) | 1 | 1 (100.00) |
| Shandong | 692 | 66 (9.54) | 6 | 6 (100.00) |
| Jiangsu | 618 | 13 (2.10) | 0 | 0 (100.00) |
| Chongqing | 576 | 0 (0.00) | 6 | 6 (100.00) |
| Sichuan | 535 | 4 (0.75) | 3 | 3 (100.00) |
| Heilongjiang | 475 | 6 (1.26) | 13 | 9 (69.20) |
| Beijing | 425 | 3 (0.71) | 8 | 3 (37.50) |
| Shanghai | 339 | 3 (0.88) | 3 | 3 (100.00) |
| Hebei | 312 | 6 (1.92) | 6 | 6 (100.00) |
| Fujian | 294 | 2 (0.68) | 1 | 1 (100.00) |
| Guangxi | 250 | 2 (0.80) | 2 | 2 (100.00) |
| Shanxi | 239 | 6 (2.51) | 1 | 1 (100.00) |
| Yunnan | 171 | 3 (1.75) | 2 | 1 (50.00) |
| Hainan | 163 | 5 (3.07) | 6 | 6 (100.00) |
| Guizhou | 144 | 2 (1.39) | 2 | 2 (100.00) |
| Tianjin | 132 | 4 (3.03) | 3 | 3 (100.00) |
| Shanxi | 131 | 2 (1.53) | 0 | 0 (100.00) |
| Liaoning | 125 | 0 (0.00) | 1 | 0 (0.00) |
| Gansu | 123 | 1 (0.81) | 2 | 2 (100.00) |
| Jilin | 92 | 1 (1.09) | 1 | 1 (100.00) |
| Xinjiang | 76 | 0 (0.00) | 3 | 3 (100.00) |
| Neimenggu | 73 | 2 (2.74) | 1 | 1 (100.00) |
| Ningxia | 75 | 0 (0.00) | 0 | 0 (100.00) |
| Hubei* | NA | NA | NA | NA |
| Total | 12698 | 275 (2.17) | 112 | 94 (83.9) |

\*Hubei Province was excluded from this study

**Appendix Table A2:** Results from unweighted logistic regression

|  | Multivariate |  |  |  |  |  |
| --- | --- | --- | --- | --- | --- | --- |
|  | OR (95% CI) | P Value | OR (95% CI) | P Value | OR (95% CI) | P Value |
| Age | 1.0 (1.0-1.1) | **0.03** | 1.0 (1.0-1.1) | **0.007** | 1.0 (1.0-1.1) | **0.05** |
| Male | 1.1 (0.6-1.7) | 0.98 | 1.1 (0.6-1.8) | 0.87 | 1.0 (0.5-1.7) | 0.89 |
| Before 01/11/2020 | 1.7 (0.9-3.1) | 0.09 | 1.6 (0.9-2.9) | 0.12 | 1.7 (0.9-3.1) | 0.09 |
| Total number of comorbidities | 1.2 (1.2-1.8) | **<0.001** | - | - | - | - |
| CHD | - | - | 3.3 (1.6-6.9) | **0.002** | 3.4 (1.7-7.4) | **0.001** |
| Cerebral infarction | - | - | - | - | 2.5 (0.9-7.0) | 0.08 |
| COPD | - | - | - | - | 2.5 (0.7-9.4) | 0.14 |
| Renal failure | - | - | - | - | 1.9 (0.5-7.0) | 0.30 |

OR=odds ratio;
CHD=coronary heart disease;
COPD=chronic obstructive pulmonary disease;
Bold: statistically significant using threshold $p < 0.05$ .

